# Therapeutic Response by Radiologic Pattern of Lung Injury in Myositis-Associated Interstitial Lung Disease: A Retrospective Cohort Study

**DOI:** 10.64898/2026.03.03.26347563

**Authors:** Thomas C. Bolig, Kevin Grudzinski, Malek Shawabkeh, Kavitha C. Selvan, Raymond J. Goodwin, Emily Olson, Bradford C. Bemiss, Nishant Parekh, Hatice Savas, Jane E. Dematte, Anthony J. Esposito

## Abstract

**Objective:** Myositis-associated interstitial lung disease (myositis-ILD) consists of two predominant radiologic patterns of lung injury—nonspecific interstitial pneumonia (NSIP) and organizing pneumonia (OP)—that oftentimes coexist. However, it remains unclear whether either is associated with clinical outcomes. We aimed to assess the therapeutic response in patients with NSIP-compared to those with OP-predominant myositis-ILD.

**Methods:** This retrospective, single-center cohort study recruited participants from the Northwestern University ILD Registry with a circulating myositis-associated antibody, ILD, and at least 6 months of follow-up while on immunomodulatory therapy during a 24-month observation period after diagnosis. Two thoracic radiologists determined the predominant radiologic pattern (NSIP or OP). The primary outcome was the absolute change in forced vital capacity (FVC) at 24 months post-diagnosis. Secondary outcomes included changes in the diffusing capacity of the lung for carbon monoxide (DLCO) and radiologic qualitative and quantitative measures of lung injury.

**Results:** Forty-one participants were included in analyses. 71% had an OP-predominant while 29% had an NSIP-predominant radiologic pattern of lung injury. Both exposure cohorts had improvement in mean absolute FVC (OP cohort = +0.18L [p=0.005], NSIP cohort = +0.24L [p=0.07]) over the 24-month observation period. The OP (p<0.05) but not the NSIP cohort (p=0.20) had an increase in DLCO. The OP cohort demonstrated improvement in the qualitative assessment of follow-up imaging (p<0.05), driven by quantitative improvement in groundglass/consolidative opacities (p=0.006). A subset of participants demonstrated features of NSIP/OP overlap and had greater baseline radiologic severity of lung injury.

**Conclusion:** Patients with circulating myositis-associated antibodies and an OP-predominant pattern of lung injury may have a more favorable response to therapy than those with NSIP. Further studies are needed to validate our findings and delineate other features cognate with these associations.

**Significance and Innovations:**

- **Radiologic phenotyping may predict therapeutic response in myositis-ILD**. This study demonstrates that an OP-predominant computed tomography (CT) pattern of lung injury is associated with greater improvement in lung function and radiologic signs of inflammation over 24 months on at least 6 months of immunomodulatory therapy compared with an NSIP-predominant pattern, suggesting that CT pattern may provide clinically meaningful prognostic information.
- **First study to integrate blinded qualitative radiologic adjudication with quantitative CT scoring in myositis-ILD**. By combining dual-radiologist review with Kazerooni quantitative scoring and longitudinal pulmonary function testing, this study offers a rigorous and multidimensional assessment of treatment response.
- **Expands risk stratification beyond antibody-based toward imaging-based phenotyping strategies**. In a heterogeneous population defined by diverse myositis-associated antibodies, this work introduces radiologic pattern as a practical and accessible framework for anticipating treatment responsiveness.
- **Provides hypothesis-generating data for precision management in myositis-ILD**. The findings support the concept that imaging-defined subgroups may exhibit differential therapeutic trajectories, laying groundwork for future multicenter studies integrating CT phenotype, antibody profile, and treatment strategy.

## INTRODUCTION

Interstitial lung disease (ILD) is a common complication of the idiopathic inflammatory myopathies (IIMs) associated with increased morbidity and mortality.^1^ The diagnosis of myositis-associated ILD (myositis-ILD) poses a challenge fueled by expansion of identified myositis-associated antibodies (MAAs) and recognition that ILD may precede muscle or skin involvement as the initial manifestation of IIMs.^2,3^ In patients with myositis-ILD, two computed tomography (CT) patterns of lung injury predominate—nonspecific interstitial pneumonia (NSIP) and organizing pneumonia (OP).^3,4^ It remains unclear whether either of these patterns are associated with clinical outcomes. This observational study examined the response to immunomodulatory therapies in two retrospective cohorts of participants with myositis-ILD: those with an NSIP-predominant pattern and those with an OP-predominant pattern of lung injury. Our findings represent an initial step toward predicting response to therapy, guiding management decisions, and generating hypotheses regarding discrete phenotypes in this patient population.

## METHODS

### Participant Data, Study Criteria, and Assignment of Exposure

Participant data were obtained from the Northwestern University (NU) ILD Registry from 01/01/2000 through 03/01/2023. The NU ILD Registry enrolled patients with confirmed ILD evaluated by the Northwestern Medicine ILD and Pulmonary Fibrosis Program who provided written informed consent for inclusion in observational research. All study procedures were approved by the NU Institutional Review Board (STU00209990).

We identified participants with ILD and circulating MAAs. The MAAs queried from the Registry were anti-PL-7, PL-12, EJ, OJ, SRP, Mi-2, U3-RNP, U2-RNP, Ku, Jo-1, TIF-1gamma, MDA5, NXP-2, PM/Scl-100, SS-A 52kD, and U1-RNP. The inclusion criteria were age > 18 years, diagnosis of ILD, an identified circulating MAA, and immunomodulatory drug use with at least six months of follow-up data on therapy nested within twenty-four months of observation from diagnosis. The exclusion criteria were an overlap connective tissue disease (CTD) or diagnosis of a separate primary CTD; clinically significant emphysema (determined by consensus review of CT imaging by thoracic radiologists); a pattern of usual interstitial pneumonia on CT imaging; and less than six months of follow up on immunomodulatory therapy.

Two thoracic radiologists reviewed chest CT imaging to assess the radiologic pattern and assign each participant to an exposure group: NSIP or OP. If a case had an overlap of NSIP and OP patterns, the radiologists determined which pattern predominated. Both radiologists independently reviewed the cases. In cases of disagreement, collaborative adjudication was performed by joint case review, and a consensus was reached. The radiologists were blinded to their previous assessments during adjudication.

### Outcomes

The primary outcome was the absolute change in forced vital capacity (FVC) over 24 months. Secondary outcomes included the absolute change in diffusing capacity of the lungs for carbon monoxide (DLCO) and qualitative and quantitative radiologic progression of ILD. Radiologic progression was assessed by the aforementioned thoracic radiologists via direct comparison of a participant’s CT at diagnosis to a follow-up CT most proximal to 24 months later.

Progression was qualitatively assessed visually and assigned categorically as “progressed”, “stable”, or “improved” on follow-up imaging. Quantitative assessment of radiologic progression was performed using the Kazerooni score, which quantifies the extent of groundglass opacity and fibrotic abnormalities on CT in patients with ILD. Each lobe is assessed for the percentage of overall involvement with scores asigned on a scale from 0 to 5 for each abnormality per lobe. Scores are summed to produce overall groudglass and fibrosis scores with higher scores indicating more extensive disease involvement. The Kazerooni score was validated by correlating CT scores with histopathologic scoring in ILD patients. ^5^

### Statistical Analysis

Fisher exact tests were performed to assess differences in categorical variables between the NSIP- and OP-exposed cohorts. Wilcoxon rank-sum tests were performed to compare changes in continuous variables. All statistical analyses were performed in R (v4.4.0)

## RESULTS

### Study Population

We identified 81 participants with circulating MAA from the NU ILD Registry. Forty-one (51%) of these participants met the inclusion/exclusion criteria and were included in the analyses. The most common reason for exclusion was an overlap CTD. The demographics were similar between the exposure groups (**Table 1**). The most common circulating MAA was anti-Jo-1 (34%), followed by anti-PL-7 (22%), anti-EJ (12%), and anti-PL-12 (10%). On review of CT imaging, 29 subjects (71%) had an OP-predominant whereas 12 (29%) had NSIP-predominant radiologic pattern of lung injury. Fifteen (37%) had an NSIP/OP overlap pattern, which were further adjudicated into a predominant pattern for exposure assignment; a majority of the overlap patterns were assigned to the OP-predominant cohort (n=11; 73%).

**Table 1.** Study Population Characteristics.

|  | **Overall**<br>**N=41** | **NSIP Group**<br>**N=12** | **OP Group**<br>**N=29** |
| --- | --- | --- | --- |
| **Age, mean (SD)** | 56 (15.7) | 53 (18.9) | 57 (14.3) |
| **BMI, mean (SD)** | 29.8 (6.6) | 30.8 (7.9) | 29.3 (5.7) |
| **<u>Sex, N (percent)</u>** |  |  |  |
| **Female** | 22 (54%) | 8(67%) | 14 (48%) |
| **Male** | 19 (46%) | 4(33%) | 15 (52%) |
| **<u>Race, N (percent)</u>** |  |  |  |
| **Asian** | 4 (10%) | 1 (8%) | 3 (10%) |
| **Asian-Indian** | 1 (2%) | 1 (8%) | 0 (0%) |
| **Black** | 11 (27%) | 3 (25%) | 8 (28% ) |
| **Caucasian** | 20 (49%) | 5 (42%) | 15 (52%) |
| **Latino** | 2 (5%) | 0 (0%) | 2 (7%) |
| **Other** | 3 (7%) | 2 (17%) | 1 (3%) |
| **<u>Smoking Status, N (percent)</u>** |  |  |  |
| **Current** | 1 (2%) | 0 (0%) | 1 (4%) |
| **Former** | 15 (37%) | 3 (25%) | 12 (41%) |
| **Never** | 25 (61) | 9 (75%) | 16 (55%) |
| **<u>Comorbidities, N (percent)</u>** |  |  |  |
| **Cardiac** | 8 (20%) | 2 (17%) | 6 (21%) |
| **CKD** | 3 (7%) | 1 (8%) | 2 (7%) |
| **GERD** | 10 (24%) | 4 (33%) | 6 (21%) |
| **Pulmonary (asthma, COPD, PH, OSA)** | 4 (10%) | 1 (8%) | 3 (10%) |
| **Malignancy** | 2 (5%) | 2 (17%) | 0 (0%) |
| **<u>Antibody, N (percent)</u>** |  |  |  |
| **Anti-EJ** | 5 (12%) | 0 (0%) | 5 (17%) |
| **Anti-Jo1** | 14 (34%) | 5 (42%) | 9 (31%) |
| **Anti-MDA5** | 1 (2%) | 0 (0%) | 1 (4%) |
| **Anti-Mi-2** | 2 (5%) | 1 (8%) | 1 (4%) |
| **Anti-OJ** | 2 (5%) | 0 (0%) | 2 (7%) |
| **Anti-PL-7** | 9 (22%) | 2 (17%) | 7 (23%) |
| **Anti-PL-12** | 4 (10%) | 1 (8%) | 3 (10%) |
| **Anti-SRP** | 3 (8%) | 2 (17%) | 1 (4%) |
| **Anti-TIF** | 1 (2%) | 1 (8%) | 0 (0%) |
| **<u>Immunomodulatory Therapy, N (percent)</u>** |  |  |  |
| **Azathioprine** | 18 (44%) | 7 (58%) | 11 (40%) |
| **Cyclophosphamide** | 3 (7%) | 1 (8%) | 2 (7%) |
| **IVIG** | 14 (34%) | 3 (25%) | 11 (38%) |
| **Mycophenolate** | 21 (51%) | 5 (42%) | 16 (55%) |
| **Prednisone** | 40 (98%) | 11 (92%) | 29 (100%) |
| **Rituximab** | 11 (27%) | 2 (17%) | 9 (31%) |
| **Tacrolimus** | 23 (56%) | 5 (42%) | 18 (62%) |
| **Tocilizumab** | 1 (2%) | 0 (0%) | 1 (4%) |
| **Follow-up PFT, months, median [interquartile range]** | 19.9 [16.4-22.3] | 20.8 [16.4-22.9] | 18.8 [16.7-21.7] |
| **Follow-up CT Chest, months, median [interquartile range]** | 17.9 [11.2-21.3] | 19.5 [14.9-23.9] | 16.0 [ 10.0-20.7] |

Abbreviations: BMI=Body Mass Index; CKD=Chronic Kidney Disease; COPD=Chronic Obstructive Pulmonary Disease; GERD=Gastroesophageal Reflux Disease; IVIG=Intravenous Immunoglobulin; OSA= Obstructive Sleep Apnea; PH=Pulmonary Hypertension; SD=standard deviation.

A variety of immunomodulatory agents were utilized during the follow-up period. These agents were typically used in succession (e.g., high-dose prednisone followed by a steroid-sparing agent) or in combination. All but one subject (98%) received prednisone. Most subjects received tacrolimus (56%) and/or mycophenolate (51%). Other immunosuppressive agents utilized were azathioprine (44%), rituximab (27%), and cyclophosphamide (7%). Intravenous immunoglobulin was administered to 34% of subjects.

No significant differences were observed in demographics, antibody distribution, or treatment between the two exposure groups (**Table 1**). Notably, two participants in the NSIP cohort had a history of malignancy compared to none in the OP cohort. One had a history of squamous cell carcinoma of the skin, and the other had breast cancer in remission. The OP group had slightly shorter median time to follow-up PFT and CT.

### Primary Outcome

Over the twenty-four month follow up period, both cohorts had an increase in mean FVC (**Figure 1A**). In the OP-exposed cohort, the mean FVC increased by an absolute change of +0.36L (p=0.006). In the NSIP-exposed cohort, the absolute change was +0.25L in the mean FVC during follow up although it did not reach statistical significance (p=0.09).

**Figure 1.**
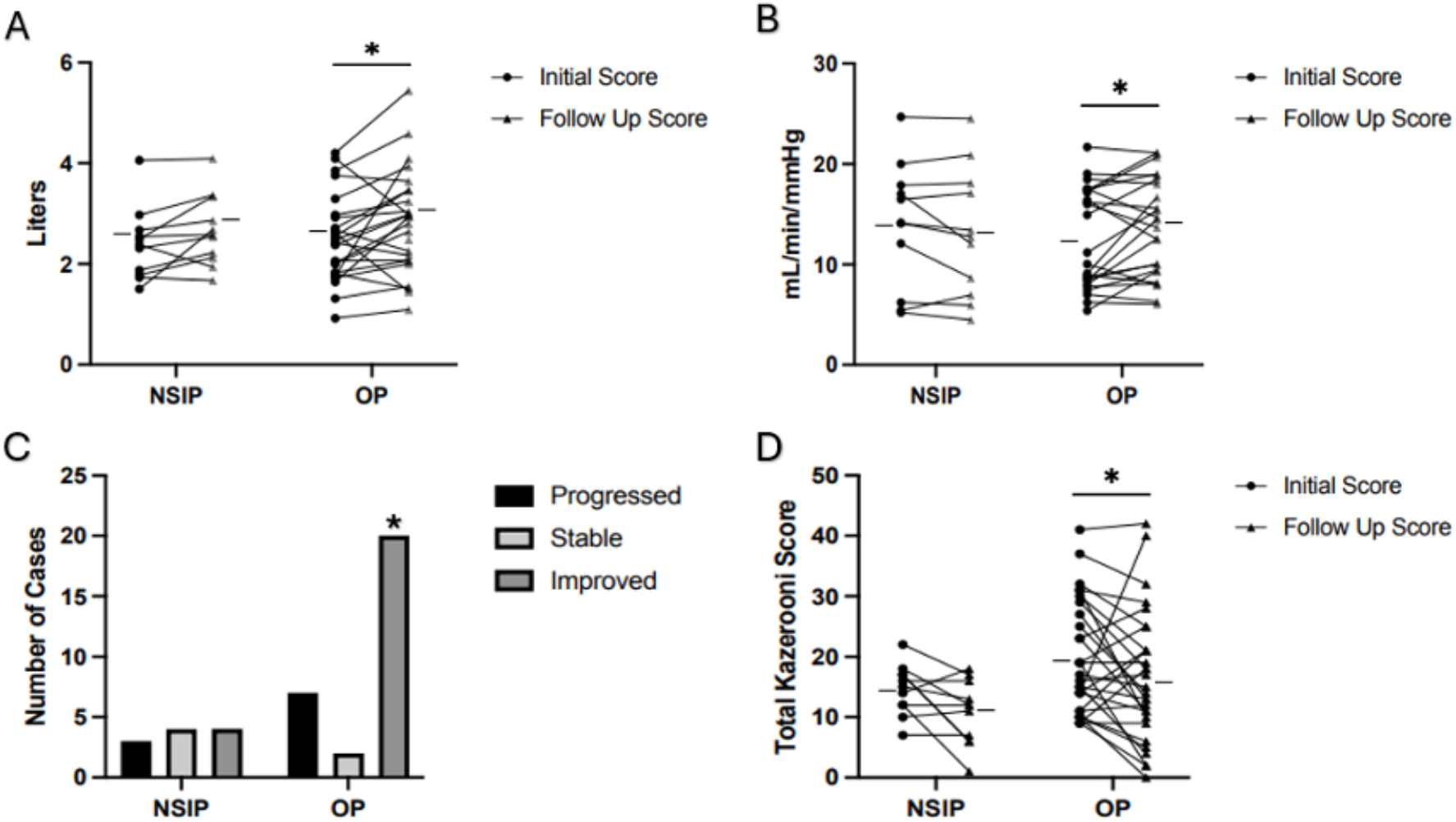
Comparisons of the NSIP- and OP-Exposed Cohorts by PFT and Radiologic Variables. (**A**) displays the change in FVC for the NSIP- and OP-exposed cohorts. Both cohorts had an improvement in FVC. The OP-exposed cohort had a statistically significant improvement in FVC. (**B**) displays the change in DLCO for both groups. The NSIP cohort had a decline in DLCO, whereas the OP cohort had an improvement. (**C**) displays the distribution of cases based on visual assessment by thoracic radiologists. The OP cohort had a statistically significant number of cases that were assessed as improved. (**D**) displays the change in the total Kazerooni score. Both groups had a decline in total Kazerooni score, indicating an improvement in the degree of lung involvement by assessing a combination of groundglass/consolidative attenuation and fibrosis. The OP cohort had a statistically significant improvement in the total Kazerooni score. * = p<0.05 — = mean value in Panels A, B, and D, displayed on sides of spaghetti plots Abbreviations: DLCO= diffusing capacity of the lung for carbon monoxide; FVC=forced vital capacity; NSIP=nonspecific interstitial pneumonia; OP= organizing pneumonia.

### Secondary Outcomes

The OP-exposed cohort had an increase in the mean DLCO for an absolute change of +1.22mL/min/mmHg (**Figure 1B**; p=0.1). Conversely, the NSIP-exposed cohort had a non-significant decrease, with an absolute change in mean DLCO of −0.75mL/min/mmHg over the 24-month observation period (p=0.4).

The OP-exposed cohort had twenty cases that demonstrated radiologic improvement (p<0.05) as visually assessed by two thoracic radiologists (**Figure 1C**). The NSIP group did not have a significant difference among the categories of “progressed”, “stable”, or “improved” lung injury on follow-up CTs.

By quantitative analysis of radiologic imaging by Kazerooni scores, the OP-exposed cohort had an initial mean total score of 19.7 that improved to 16.7 (**Figure 1D**; p=0.04). The fibrosis subscore worsened from 6.1 to 8.5 (p=0.02) whereas the groundglass/consolidation subscore significantly improved from 13.5 to 8.2 (p=0.0002). The NSIP-exposed cohort had an initial mean total score of 14.5 that improved to 10.8, which approached statistical significance (p=0.06). The mean change in the fibrosis subscore was 8.4 to 7.7 (p=0.36), and the mean change in groundglass/consolidation subscore was 6.2 to 3.1 (p=0.06).

#### Exploratory Analysis of NSIP-OP Overlap

Given NSIP/OP overlap has been previously associated with progression of ILD,^6^ we performed an exploratory analysis of participants with overlap features (n=15). As expected, those with overlap had significantly greater radiologic involvement at baseline, with a higher mean total Kazerooni score compared to those without overlap (**Table 2**; 25.0 vs. 14.5, p=0.0001). Both overlap and non-overlap subgroups demonstrated improvement in FVC over follow-up, without significant changes in DLCO. Improvement in total Kazerooni score was observed in the overlap subgroup (p=0.004), largely driven by reduction in groundglass/consolidation. Rates of hospitalization and supplemental oxygen use were numerically higher in the overlap subgroup (47% vs. 31% and 27% vs. 15%, respectively), though these differences were not statistically significant.

**Table 2.** Characteristics and Outcomes of Cohorts with and without NSIP/OP Overlap.

|  | NSIP/OP Overlap<br>(N=15) | NSIP or OP Only<br>(N=26) |
| --- | --- | --- |
| Predominant Disease Pattern |  |  |
| NSIP, N (percent) | 4 (27) | 8 (31) |
| OP, N (percent) | 11 (73) | 18 (69) |
| FVC (Liters) |  |  |
| Initial, mean (SD) | 2.5 (0.9) | 2.5 (0.8) |
| Follow Up, mean (SD) | 2.7 (1.1) | 3.0 (0.8) |
| DLCO (mL/min/mmHg) |  |  |
| Initial, mean (SD) | 12.05 (6.62) | 13.50 (4.46) |
| Follow Up, mean (SD) | 13.24 (6.08) | 13.71 (4.92) |
| Kazerooni Score |  |  |
| Initial, mean (SD) | 24.9 (8.5) | 14.28 (5.5) |
| Follow Up, mean (SD) | 17.8 (11.1) | 13.40 (8.9) |
| Hospitalizations*, N (percent) | 7 (47) | 8 (31) |
| Supplemental O<sub>2</sub><sup>†</sup>, N (percent) | 4 (27) | 4 (15) |

Abbreviations: DLCO = diffusing capacity for carbon monoxide; FVC = forced vital capacity; NSIP=non-specific interstitial pneumonia; OP= organizing pneumonia.
\*Hospitalizations were defined as the number of inpatient admissions during the follow up period.
<sup>†</sup>Supplemental O<sub>2</sub> is the number of times patients either initiated supplemental oxygen therapy or had an increase in their baseline oxygen requirements after an outpatient assessment.

## DISCUSSION

In this observational cohort, we found that participants with circulating MAAs and an OP-predominant pattern of lung injury demonstrated greater improvement in lung function and radiologic signs of inflammation over 24 months compared with those with an NSIP-predominant pattern. These findings suggest that radiologic patterns may provide clinically meaningful information regarding therapeutic responsiveness in myositis-ILD. To our knowledge, this is among the first studies to evaluate outcomes in myositis-ILD stratified by predominant radiologic pattern of lung injury while incorporating both physiologic and quantitative imaging measures. The use of blinded dual-radiologist review and quantitative CT scoring strengthens the internal validity of our findings. These data suggest that radiologic phenotyping may represent an accessible and clinically useful approach to anticipating therapeutic response in this heterogeneous population.

Assessing prognosis in patients with myositis-associated ILD remains challenging, with novel approaches to phenotyping and risk stratification under active investigation.^3,7^ This is underscored by the rarity of the disease in the general population and the lack of large clinical studies to guide assessment. Participants with an OP-predominant pattern experienced a significant increase in FVC, along with qualitative and quantitative radiologic improvement. In contrast, those with an NSIP-predominant pattern did not demonstrate statistically significant improvement in FVC or qualitative radiologic outcomes, although quantitative measures suggested improvement in the groundglass/consolidation component of lung injury. Together, these findings suggest that OP-predominant disease may demonstrate a more pronounced short-term physiologic response to immunomodulatory therapy compared with NSIP-predominant disease, although both patterns likely reflect inflammatory lung injury with varying degrees of reversibility.

The observed differences between radiologic patterns may be explained by the relative reversibility of radiologic signs of active lung inflammation with immunomodulatory therapy. The OP-exposed cohort had a higher baseline groundglass/consolidation burden, and improvement in this component appeared to drive both qualitative radiologic improvement and overall Kazerooni score reduction. This pattern is biologically plausible, as OP is typically characterized by inflammatory consolidation that may respond favorably to immunomodulation. In contrast, NSIP may encompass a greater fibrotic component, which may be less responsive to anti-inflammatory therapy and therefore associated with more modest physiologic gains.

Although a substantial subset of our cohort exhibited NSIP/OP overlap features, evaluation of overlap status was not a primary objective of this study. Prior literature in CTD-ILD has suggested that NSIP/OP overlap may be associated with worse outcomes.^6^ Consistent with these reports, participants with overlap in our cohort demonstrated significantly greater baseline radiologic involvement. However, subgroup sizes were small, and exploratory analyses showed only numerically higher rates of hospitalization and supplemental oxygen use without statistically significant differences. While some improvement in radiologic inflammation was observed in the overlap subgroup, these findings should be interpreted cautiously. Accordingly, our observations regarding NSIP/OP overlap are hypothesis-generating and warrant validation in larger cohorts.

Several limitations merit consideration. This retrospective single-center study is subject to referral and treatment selection biases, which may limit generalizability. Therapeutic regimens were heterogeneous, and we were unable to account for differences in response attributable to specific MAAs or individual immunomodulatory agents. Additionally, some participants had their first follow-up pulmonary function test after initiation of immunomodulation during an index hospitalization, potentially attenuating the magnitude of observed physiologic change. The relatively small sample size also limits statistical power, particularly for subgroup analyses. Nevertheless, given the rarity of myositis-ILD, our cohort size is comparable to many existing studies in this field that have impacted clinical management.^4,10,11^

In conclusion, among patients with myositis-ILD, an OP-predominant radiologic pattern was associated with greater improvement in lung function and radiologic signs of inflammation over 24 months compared with an NSIP-predominant pattern. These findings suggest that CT-based phenotyping may provide prognostic and therapeutic insight in myositis-ILD. Larger, multicenter studies are needed to validate these observations and to determine whether integration of radiologic pattern, antibody profile, and treatment strategy can more precisely guide management in this complex disease.

## Funding statement

This study was funded by NIH/NHLBI T32HL076139 (T.C.B.), NIH/NHLBI L30HL149048 (A.J.E.), and the Pulmonary Fibrosis Foundation Scholars program (A.J.E.).

## Conflict of interest statement

The authors declare that they have no known competing financial interests or personal relationships that could have appeared to influence the work reported in this paper.

## Data availability statement

The data utilized for analyses in this manuscript will be shared upon reasonable request to the corresponding author.

## Notes

### Competing Interest Statement

The authors have declared no competing interest.

### Author Declarations

All study procedures were approved by the Northwestern University Institutional Review Board (STU00209990).

